# Intestinal function and transit associate with gut microbiota dysbiosis in cystic fibrosis

**DOI:** 10.1101/2021.08.24.21262265

**Authors:** Ryan Marsh, Helen Gavillet, Liam Hanson, Christabella Ng, Mandisa Mitchell-Whyte, Giles Major, Alan R Smyth, Damian Rivett, Christopher van der Gast

## Abstract

**Background:** Most people with cystic fibrosis (pwCF) suffer from gastrointestinal symptoms and are at risk of gut complications. Gut microbiota dysbiosis is apparent within the CF population across all age groups, with evidence linking dysbiosis to intestinal inflammation and other markers of health. This pilot study aimed to investigate the potential relationships between the gut microbiota and gastrointestinal physiology, transit, and health.

**Study Design:** Faecal samples from 10 pwCF and matched controls were subject to 16S rRNA sequencing. Results were combined with clinical metadata and MRI metrics of gut function to investigate relationships.

**Results:** pwCF had significantly reduced microbiota diversity compared to controls. Microbiota compositions were significantly different, suggesting remodelling of core and rarer satellite taxa in CF. Dissimilarity between groups was driven by a variety of taxa, including *Escherichia coli, Bacteroides spp*., *Clostridium spp*., and *Faecalibacterium prausnitzii*. The core taxa were explained primarily by CF disease, whilst the satellite taxa were associated with pulmonary antibiotic usage, CF disease, and gut function metrics. Species-specific ordination biplots revealed relationships between taxa and the clinical or MRI-based variables observed.

**Conclusions:** Alterations in gut function and transit resultant of CF disease are associated with the gut microbiota composition, notably the satellite taxa. Delayed transit in the small intestine might allow for the expansion of satellite taxa resulting in potential downstream consequences for core community function in the colon.

**Highlights:**

- Faecal microbiota significantly differs between pwCF and healthy controls
- Key SCFA producers contributed to microbiota dissimilarity between groups
- Pulmonary antibiotic treatment heavily impacted gut microbiota
- Intestinal physiology and transit impacted satellite microbiota composition

## 1. Introduction

Cystic fibrosis (CF) associated respiratory infections are the major cause of disease morbidity and mortality. However, a number of gastrointestinal (GI) problems may also arise, limiting the quality of life, including meconium ileus at birth, distal intestinal obstruction syndrome, small intestinal bacterial overgrowth (SIBO), increased risk of malignancy, and intestinal inflammation [1,2]. It is therefore unsurprising that people with CF experience persistent GI symptoms [3,4] with “how can we relieve gastrointestinal symptoms in people with CF?” a top priority question for research [5].

Microbial dysbiosis at the site of the GI tract in CF patients has been described, with changes evident from birth through to adulthood [6–8]. Moreover, the extent of this divergence from healthy microbiota, initially due to loss of cystic fibrosis transmembrane conductance regulator (CFTR) function [9], is further compounded by routine treatment with broad spectrum antibiotics [10]. The reshaping of the gut microbiota may have functional consequences that could further impact on patients. These include the reduction of taxa associated with the production of short-chain fatty acids (SCFAs) which play key roles in modulating local inflammatory responses and promoting gut epithelial barrier integrity [11–13]. Furthermore, studies of microbiota dysbiosis in CF have demonstrated its relationship with intestinal inflammation [14], intestinal lesions [15], and increased gene expression relating to intestinal cancers [16]. Whilst many of these clinical parameters have ties to gut microbiota changes, they remain understudied exclusively past childhood despite advances in less invasive approaches to investigate CF gut physiology and function [17]. Our group has recently published on the use of magnetic resonance imaging (MRI) to assess gut transit time, along with other parameters, in adolescents and adults [18].

In this pilot study, we linked those MRI physiology metrics and clinical metadata directly to high-throughput amplicon sequencing data identifying constituent members of the gut microbiota, to explore the relationships between microbial dysbiosis, intestinal function and clinical state.

## 2. Materials and methods

### 2.1. Study participants and design

Twelve people with CF, homozygous for p.Phe508del along with 12 healthy controls, matched by age and gender, were recruited from Nottingham University Hospitals NHS Trust. Participants were asked to provide stool samples when attending for MRI scanning, with the study design and MRI protocols described previously [18]. A patient clinical features were also recorded upon visitation (Table 1), including a three-day food diary preceding sample collection (Table S1). Further descriptive statistics of the study population can be found in the Supplementary Materials, including MRI metrics (Table S2), and summary statistics on diet (Tables S3-S6). Faecal samples were only obtained from ten individuals in each group. Written informed consent, or parental consent and assent for paediatric participants, was obtained from all participants. Study approval was obtained from the West Midlands Coventry and Warwickshire Research Ethics Committee (18/WM/0242). All stool samples obtained were immediately stored at -80°C prior to DNA extraction to reduce changes before downstream community analysis [19].

**Table 1.** Clincial characteristics of study participants.

| Study I.D | Sex | Age (Years) | Group | Pancreatic Status | Calprotectin (µg/g) | FEV1% | BMI | Antibiotic Usage |  |  |  |  |
| --- | --- | --- | --- | --- | --- | --- | --- | --- | --- | --- | --- | --- |
|  |  |  |  |  |  |  |  | P | A | M | β | S |
| 365 | M | 12-16 | CF | PI | 4.22 | 87 | 16.18 | - | - | - | + | - |
| 431 | M | 12-16 | HC | PS | 2.44 | - | 17.95 | - | - | - | - | - |
| 128 | M | 12-16 | CF | PI | 27.59 | 97 | 17.72 | + | - | - | - | - |
| 296* | M | 12-16 | HC | PS | - | - | 23.44 | - | - | - | - | - |
| 643 | M | 12-16 | CF | PI | 9.77 | 90 | 21.83 | - | - | + | - | - |
| 159 | M | 12-16 | HC | PS | 2.72 | - | 23.49 | - | - | - | - | - |
| 297 | M | 12-16 | CF | PI | 27.61 | 126 | 20.83 | - | - | - | - | - |
| 947* | M | 12-16 | HC | PS | - | - | 20.94 | - | - | - | - | - |
| 617 | F | 12-16 | CF | PI | 21.15 | 72 | 18.42 | - | - | + | + | + |
| 964 | F | 12-16 | HC | PS | 12.71 | - | 19.15 | - | - | - | - | - |
| 167 | M | 17-21 | CF | PI | 7.37 | 99 | 20.63 | - | - | - | - | - |
| 673 | M | 17-21 | HC | PS | 0.94 | - | 20.34 | - | - | - | - | - |
| 279 | F | 17-21 | CF | PI | 27.32 | 66 | 20.87 | - | - | + | - | - |
| 205 | F | 17-21 | HC | PS | 3.84 | - | 31.91 | - | - | - | - | - |
| 596 | F | 17-21 | CF | PI | 14.05 | 61 | 21.91 | - | - | + | - | - |
| 152 | F | 17-21 | HC | PS | 4.22 | - | 21.26 | - | - | - | - | - |
| 610* | M | 23-27 | CF | PI | - | 66 | 18.64 | - | + | + | - | - |
| 548 | M | 23-27 | HC | PS | 3.56 | - | 24.49 | - | - | - | - | - |
| 619* | F | 23-27 | CF | PI | - | 60 | 19.27 | - | - | - | - | - |
| 501 | F | 23-27 | HC | PS | 7.19 | - | 28.66 | - | - | - | - | - |
| 259 | M | 28-32 | CF | PI | 28.30 | 61 | 20.21 | - | - | + | + | - |
| 986 | M | 28-32 | HC | PS | 4.96 | - | 22.64 | - | - | - | - | - |
| 681 | F | 33-37 | CF | PI | 11.79 | 88 | 21.71 | + | - | + | - | - |
| 749 | F | 33-37 | HC | PS | 3.00 | - | 19.57 | - | - | - | - | - |

Subjects marked with an asterisk\* indicate those who failed to produce a stool sample for subsequent metagenomic and metabolomic analysis and thus were excluded from downstream analyses. All participants with CF had the gene mutation p.Phe508del/p.Phe508del, with pancreatic insufficiency but no CF-related diabetes. For antibiotic usage, '+' indicates routine administration of the given antibiotic class prior to sampling. Abbreviations: FEV1 – Percent predicted forced expiratory volume in 1 second, BMI – Body mass index, P – Polymyxin, A – Aminoglycoside, M – Macrolide, β – β-lactam, S – Sulfonamide. Asterisks denote participants who did not provide any stool samples upon visitation, and thus were excluded from downstream microbiota analysis.

### 2.2. Targeted amplicon sequencing

DNA from dead or damaged cells, as well as extracellular DNA was excluded from analysis via cross-linking with propidium monoazide (PMA) prior to DNA extraction, as previously described [20]. Next, cellular pellets resuspended in PBS were loaded into the ZYMO Quick-DNA Fecal/Soil Microbe Miniprep Kit (Cambridge Bioscience, Cambridge, UK) as per manufacturer’s instructions, with the following amendments: ZR BashingBead Lysis Tubes were replaced with standard 1.5 mL Eppendorf tubes loaded with ZYMO Beads for mechanical homogenisation with the use of a Retsch Mixer Mill MM 400 (Retsch, Haan, Germany). Samples were homogenised for 2 minutes at 17.5/s frequency. Following DNA extraction, approximately 20 ng of template DNA was then amplified using Q5 high-fidelity DNA polymerase (New England Biolabs, Hitchin, UK) using a paired-end sequencing approach targeting the bacterial 16S rRNA gene region (V4-V5). Primers and PCR conditions can be found in the Supplementary Materials. Pooled barcoded amplicon libraries were sequenced on the Illumina MiSeq platform (V3 Chemistry).

### 2.3. Sequence processing and analysis

Sequence processing and data analysis were initially carried out in R (Version 4.0.1), utilising the package DADA2 [21]. The full protocol is detailed in the Supplementary Materials. Raw sequence data reported in this study has been deposited in the European Nucleotide Archive under the study accession number PRJEB44071.

### 2.4. Faecal Calprotectin

Stool was extracted for downstream assays using the ScheBo® Master Quick-Prep (ScheBo Biotech, Giessen, Germany), according to the manufacturer instructions. Faecal calprotectin was analysed using the Bühlmann fCAL ELISA (Bühlmann Laboratories Aktiengesellschaft, Schonenbuch, Switzerland), according to the manufacturer’s protocol.

### 2.5. Statistical Analysis

Regression analysis, including calculated coefficients of determination (*r*^2^), degrees of freedom (df), *F*-statistic and significance values (*P*) were calculated using XLSTAT v2021.1.1 (Addinsoft, Paris, France). Fisher’s alpha index of diversity and the Bray-Curtis index of similarity were calculated using PAST v3.21 [22]. Significant differences in microbiota diversity were determined using Kruskal-Wallis performed using XLSTAT. Analysis of similarities (ANOSIM) with Bonferroni correction was used to test for significance in microbiota composition and was performed in PAST. Similarity of percentages (SIMPER) analysis, to determine which taxa contributed most to compositional differences between groups, was performed in PAST.

Redundancy analysis (RDA), was performed in CANOCO v5 [23]. Following the determination of clinical variables significantly explanatory for microbiome composition, RDA biplots with these variables were plotted in PAST v3.21. Statistical significance for all tests was deemed at the *p* ≤ 0.05 level. Supplementary information, including metadata, are available at figshare.com under https://doi.org/10.6084/m9.figshare.15073797.v1 and https://doi.org/10.6084/m9.figshare.15073899.v1

## 3. Results

For both healthy control and CF groups, bacterial taxa were partitioned into core or satellite based on their prevalence and relative abundance as depicted in (Fig. 1). Within the healthy control group, 30 taxa were core constituting 60.5 % of the total abundance, with the remainder accounted for by 386 satellite taxa. In the CF group, 22 core taxa represented 34.7 % of the abundance, with 323 satellite taxa constituting the remainder. Core taxa are listed in Table S7. The whole, core, and satellite microbiota demonstrated similar patterns in diversity, whereby there was significantly reduced diversity in the CF group (Fig. 2A, Table S8).

**Figure 1.**
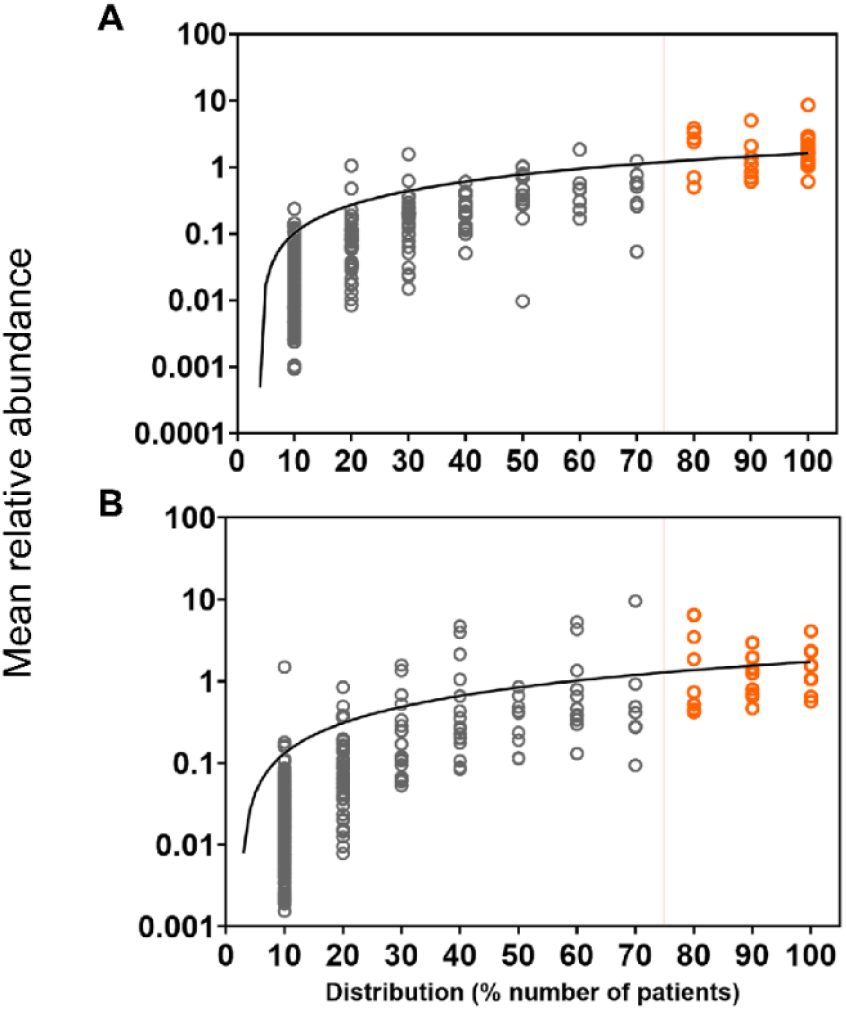
Distribution and abundance of bacterial taxa across different sample groups. (A) Healthy control. (B) Cystic fibrosis. Given is the percentage number of patient stool samples each bacterial taxon was observed to be distributed across, plotted against the mean percentage abundance across those samples. Core taxa are defined as those that fall within the upper quartile of distribution (orange circles), and satellite taxa (grey circles) defined as those that do not, separated by the red vertical line at 75% distribution. Distribution-abundance relationship regression statistics: (a) *r*^2^ = 0.50, *F*_1,414_ = 407.3, *P* < 0.0001; (b) *r*^2^ = 0.29, *F*_1,343_ = 137.3, *P* < 0.0001. Core taxa are listed in Table S7.

**Figure 2.**
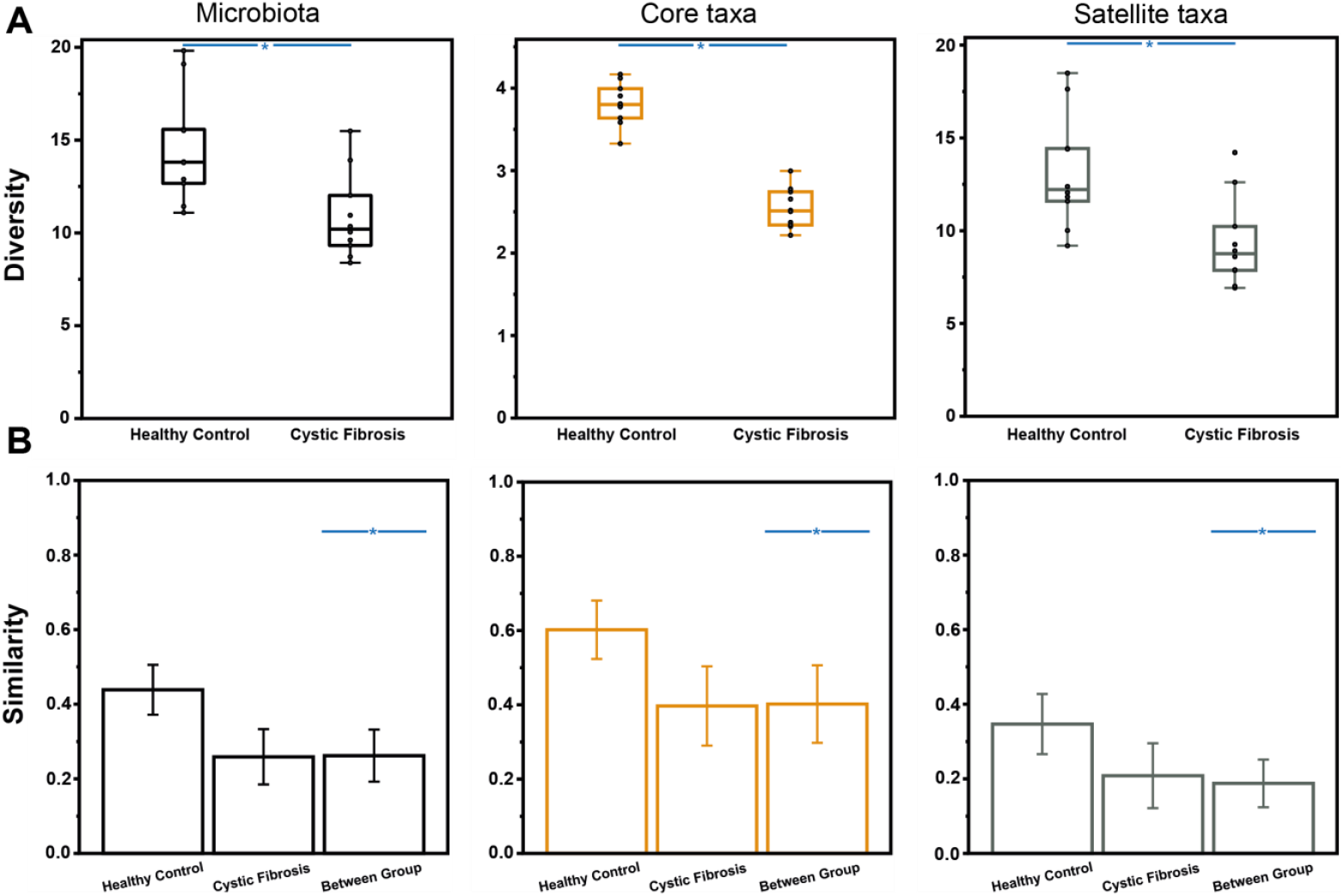
Microbiome diversity and similarity compared across healthy controls and cystic fibrosis samples. Whole microbiota (black plots) and partitioned data into core (orange plots) and satellite taxa (grey plots) are given. (**A**) Differences in Fisher’s alpha index of diversity between healthy controls and cystic fibrosis samples. Black circles indicate individual patient data. Error bars represent 1.5 times inter-quartile range (IQR). Asterisks between groups denote a significant difference in diversity following use of Kruskal-Wallis tests (*P* < 0.001). Summary statistics are provided in Table S8. (**B**) Microbiome variation measured within and between sampling groups, utilising the Bray-Curtis index of similarity. Error bars represent standard deviation of the mean. Asterisks indicate significant differences between sampling groups following the use of one-way ANOSIM testing (*P* < 0.001). Summary statistics are provided in Table S9.

Within-group core microbiota similarity was higher within the healthy control group, with a mean similarity (± SD) of 0.60 ± 0.08 compared to 0.40 ± 0.11 for the CF group (Fig. 2B). As expected, satellite taxa similarity within groups was much lower than for the core but was also significantly reduced in CF compared to controls, at 0.35 ± 0.08 and 0.21 ± 0.09 for the healthy control and CF group respectively. ANOSIM testing determined the whole microbiota, core, and satellite taxa of the CF group were significantly different in composition compared to healthy controls (Fig. 2B, Table S9). SIMPER analysis was implemented to reveal which taxa were responsible for driving this dissimilarity (Table 2). Of the taxa contributing to > 50% of the differences between healthy control and CF groups, those within the genus *Bacteroides* were represented most. *Escherichia coli* contributed most towards the differences between groups, despite satellite status, followed by *Bacteroides* sp. (OTU 3), *Clostridium* sp. (OTU 5), *Faecalibacterium prausnitzii*, and *Bacteroides fragilis*.

**Table 2.** Similarity of percentage (SIMPER) analysis of microbiota dissimilarity (Bray-Curtis) between Healthy Control (HC) and Cystic Fibrosis (CF) stool samples.

| Taxa | %Relative abundance |  | Av. Dissimilarity | % Contribution | Cumulative % |
| --- | --- | --- | --- | --- | --- |
|  | Mean HC | Mean CF |  |  |  |
| *Escherichia coli* | 1.84 | 9.54 | 4.72 | 6.39 | 6.39 |
| *Bacteroides 3* | 3.84 | 4.69 | 3.36 | 4.55 | 10.94 |
| *Clostridium 5* | 0.77 | 6.44 | 3.09 | 4.18 | 15.13 |
| *Faecalibacterium prausnitzii* | 8.56 | 2.95 | 2.99 | 4.05 | 19.18 |
| *Bacteroides fragilis* | 1.02 | 5.29 | 2.75 | 3.73 | 22.90 |
| *Bacteroides dorei* | 3.32 | 4.31 | 2.52 | 3.42 | 26.32 |
| *Eubacterium rectale* | 5.03 | 1.35 | 2.18 | 2.95 | 29.27 |
| *Romboutsia timonensis* | 1.24 | 3.95 | 2.15 | 2.91 | 32.18 |
| *Bacteroides uniformis* | 2.72 | 4.09 | 1.62 | 2.20 | 34.38 |
| *Dialister invisus* | 1.00 | 3.45 | 1.61 | 2.19 | 36.57 |
| *Bacteroides vulgatus* | 2.37 | 2.14 | 1.56 | 2.11 | 38.68 |
| *Ruminococcus bromii* | 2.69 | 0.42 | 1.24 | 1.68 | 40.36 |
| *Alistipes putredinis* | 2.08 | 0.06 | 1.02 | 1.38 | 41.74 |
| *Bacteroides coprocola* | 1.56 | 0.92 | 1.01 | 1.37 | 43.11 |
| *Fusicatenibacter saccharivorans* | 2.62 | 0.8 | 1.00 | 1.36 | 44.47 |
| *Streptococcus 18* | 0.26 | 1.95 | 0.88 | 1.19 | 45.66 |
| *Blautia luti* | 2.93 | 2.31 | 0.86 | 1.16 | 46.82 |
| *Oscillibacter ruminantium* | 1.90 | 0.27 | 0.84 | 1.14 | 47.96 |
| *Clostridium perfringens* | 0.00 | 1.58 | 0.79 | 1.07 | 49.03 |
| *Parabacteroides distasonis* | 1.38 | 1.85 | 0.77 | 1.05 | 50.08 |

Taxa identified as core are highlighted in orange, whereas satellite taxa are highlighted in grey. Mean relative abundance (%) is also provided for each group. Percentage contribution is the mean contribution divided by the mean dissimilarity across samples (73.79%). Cumulative percent does not equal 100% as the list is not exhaustive. Given the sequencing length of 16S gene regions, taxon identification should be considered putative.

Redundancy analysis (RDA) was used to relate variability in microbiota composition to associated MRI metrics and clinical factors (Table 3). Pulmonary antibiotics and CF disease significantly explained the most variance across the whole and satellite microbiota. Measurements of intestinal transit and function contributed to the whole microbiota variance, albeit to a lesser extent, with variation in OCTT and SWBC also contributing to satellite taxa variance alongside faecal calprotectin levels. In the core taxa analysis, the presence of CF disease was the dominant factor in significantly explaining the compositional variability, followed by sex and body mass index (BMI).

**Table 3.** Redundancy analysis to explain percent variation in whole microbiota, core taxa and satellite taxa between all subjects from significant clinical variables measured.

|  | **Microbiota** |  |  | **Core taxa** |  |  | **Satellite taxa** |  |  |
| --- | --- | --- | --- | --- | --- | --- | --- | --- | --- |
|  | Var. Exp (%) | pseudo-*F* | *P* (adj) | Var. Exp (%) | pseudo-*F* | *P* (adj) | Var. Exp (%) | pseudo-*F* | *P* (adj) |
| Antibiotics | 21.5 | 5.4 | 0.002 |  |  |  | 27.1 | 7.3 | 0.002 |
| BMI |  |  |  | 7.0 | 2.0 | 0.042 |  |  |  |
| Calprotectin |  |  |  |  |  |  | 5.9 | 1.8 | 0.050 |
| CF Disease | 10.9 | 2.2 | 0.002 | 28.9 | 7.3 | 0.002 | 10.3 | 2.1 | 0.006 |
| Colon Fasting Vol. | 7.5 | 2.0 | 0.016 |  |  |  |  |  |  |
| OCTT | 7.4 | 2.1 | 0.012 |  |  |  | 6.7 | 1.9 | 0.046 |
| SBWC | 5.6 | 1.7 | 0.048 |  |  |  | 7.2 | 2.4 | 0.048 |
| Sex |  |  |  | 7.9 | 2.1 | 0.010 |  |  |  |
| **Total** | **52.9** |  |  | **43.8** |  |  | **57.2** |  |  |

Var. Exp (%) represents the percentage of the microbiota variation explained by a given parameter within the redundancy analysis model. *P* (adj) is the adjusted significance value following false discovery rate correction. Antibiotics is the presence/absence of recurrent antibiotic regimes for a given patient. BMI – Body mass index, Colon Fasting Vol – Colon volume at baseline corrected for body surface area, OCTT – Oro-caecal transit time, Antibiotics, SBWC – Small bowel water content corrected for body surface area.

A species redundancy analysis biplot (RDA) was constructed to investigate how significant clinical variables from the whole microbiota direct ordination approach explained the relative abundance of taxa from the SIMPER analysis (Fig. 3). Certain taxa grouped away from many of the significant clinical variables shown in a similar manner. This effect was most pronounced for *F. prausnitzii, Eubacterium rectale* and *Ruminoccocus bromii*. A combination of clinical factors, including CF disease, increased fasting colonic volume, increased SBWC and prolonged OCTT, explained the variance observed in relative *E. coli* abundance, whilst a more modest effect was observed towards *Streptococcus* sp. (OTU 18), *Dialister invisus, Clostridium perfringens* and *Romboutsia timonensis*. Species of *Bacteroides*, which was the most common genus within the top-contributing SIMPER analysis, were explained by the clinical variables to high variability.

**Fig. 3.**
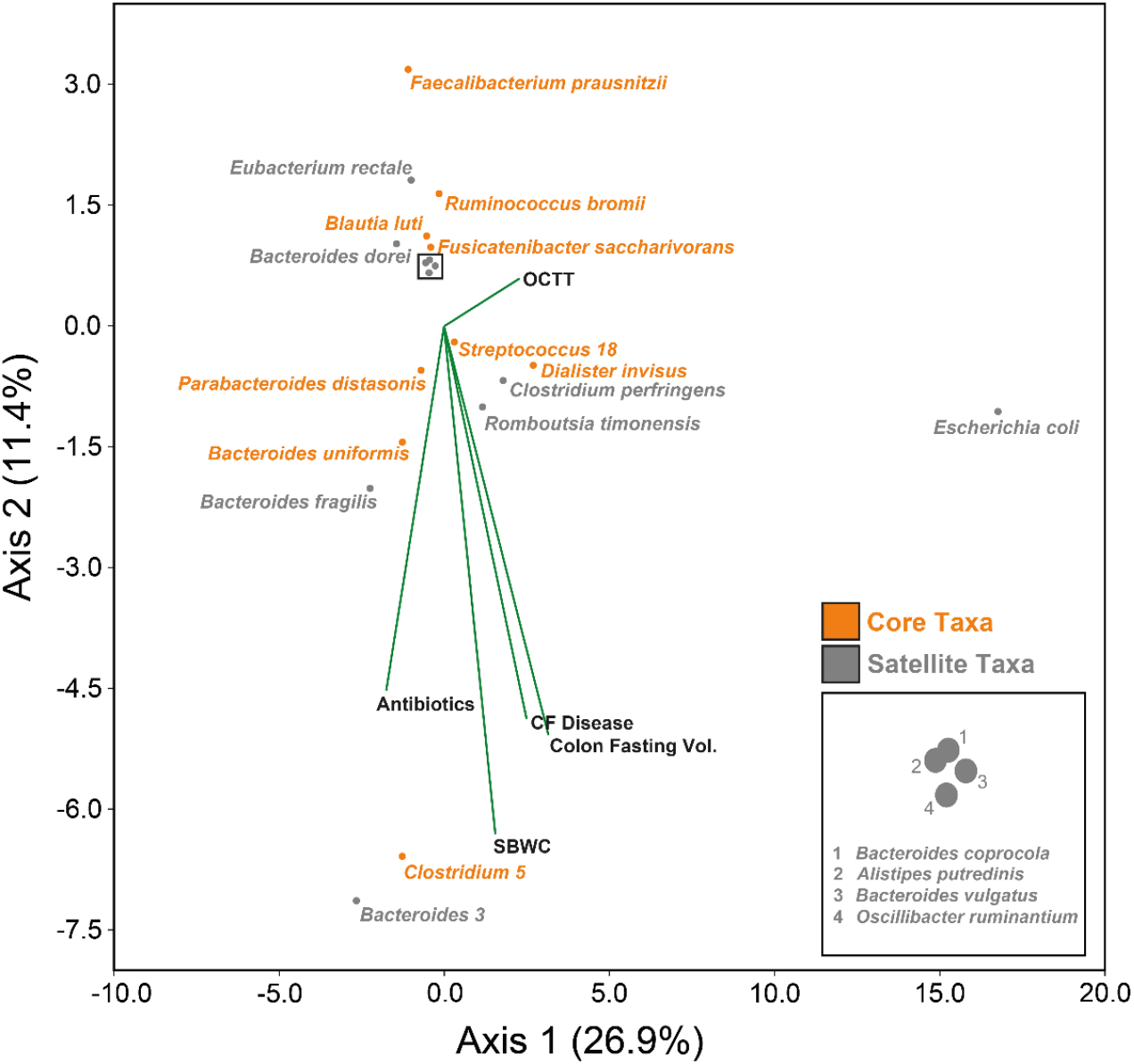
Redundancy analysis species biplots for whole microbiota. The 20 taxa contributing most to the dissimilarity (cumulatively > 50%) between healthy and cystic fibrosis groups from the SIMPER analysis (Table 2) are shown independently of the total number of ASVs identified (345). Orange circles represent core taxa within the CF group, whilst grey circles denote satellite taxa. Biplot lines depict clinical variables that significantly account for the total variation in taxa relative abundance within the whole microbiota analysis at the *p* ≤ 0.05 level as seen in Table 3, with species plots indicating the strength of explanation provided by the given clinical variables. ‘OCTT’ – Oro-caecal transit time, ‘SBWC’ – Small bowel water content corrected for body surface area, Colon Fasting Volume corrected for body surface area, CF disease. The percentage of microbiome variation explained by each axis is given in parentheses.

## 4. Discussion

In this pilot study, we investigated the relationships between clinical factors, MRI markers of GI function and the composition of faecal bacterial microbiota. We have shown that it is possible to partition the gut microbiota into core and satellite taxa to investigate potential community functions and relationships, with the notion that the core constituents contribute to the majority of functionality exhibited by the community [20,24]. As to be expected, the core taxa made up most of the abundance within the healthy control group. Whilst many taxa were also commonly represented in the CF group, the latter was dominated in abundance by the satellite taxa. Our findings of reduced diversity across the whole, core, and satellite microbiota are in agreement with previous findings described within the CF gut [7,8,10]. Along with reduced within group similarity in CF compared to healthy controls across all microbiota partitions, this suggests a perturbed community harbouring greater instability, less subsequent resilience, and inherent challenges to the colonisation and establishment of normal commensals. CF associated factors such as varied antibiotic usage will contribute to this reduced similarity, further augmented by the wide age range of pwCF within this study and variation across lifestyle factors. The combination of the aforementioned may elicit stochastic community disruption and increased inter-individual variation as observed across other mammalian microbiomes [25].

At the surface, a reduction in the number of taxa labelled as core within the CF group hinted at perturbation and restructuring, further evidenced by the occurrence of taxa exclusively core to this group. This included species of *Streptococcus, Pseudomonas, Veillonella*, and *Enterococcus*, all of which were significantly more abundant in the CF group (Table S7), and of which are implicated in both CF lung and gut microbiomes [8,9,24,26–28]. The concept of the “gut-lung axis” in CF arises from the direct translocation of the respiratory microbiota from sputum swallowing to the gut [29], but also the emergence of species in the gut prior to the respiratory environment [27]. This apparent bidirectionality is further supported by the administration of oral probiotics to decrease pulmonary exacerbations in CF [30]. Aside from sputum swallowing, the increase in *Streptococcus* and *Veillonella* here could reflect an increased availability of simple carbohydrates from the observed dysmotility of the gut [18]. *Streptococci* are well equipped with numerous genes for rapid carbohydrate degradation in an environment usually fluctuating in substrate availability, with fermentation-derived lactic acid supporting the expansion of *Veillonella* species in the small intestine [31].

*E. coli* contributed most to the dissimilarity between healthy and CF groups despite maintaining satellite status throughout both the healthy and CF groups, seemingly resultant of the wide age range of our study participants, of which the higher relative abundances were observed in the younger adolescent patients (Table 2). In childhood studies, a significantly higher relative abundance of *Proteobacteria* is often reported in relation to dysbiosis, with *E. coli* abundance associating with poor growth outcomes and intestinal inflammation [32–34]. Other notable taxa contributing to the dissimilarity observed between groups encompassed a variety of key species associated with SCFA production in the colon. This included *F. prausnitizii* and *E. rectale*, both of which were significantly decreased in abundance within the CF group, but also *R. bromii* and *B. luti*. These taxa have all been previously reported to decrease in the CF gut [8,26,35] alongside other inflammatory conditions [36]. There were also notable contributions to the dissimilarity between groups by *Clostridium* sp. (OTU 5) (significant difference in relative abundance) and *D. invisu*s (not significant). *Clostridium* OTU 5 aligned exclusively with cluster I members at the 97% threshold, of whom demonstrate the capacity to generate lactate, acetate, propionate, and butyrate via carbohydrate fermentation [37], whilst *D. invisu*s is an intermediary fermenter capable of both acetate and propionate production. This may lend support to the theory that alternate species can retain some functional redundancy in the presence of perturbation to the local community in the CF gut [38].

Variance across the whole microbiota and satellite taxa was significantly explained by the use of antibiotics (Table 3), of which most pwCF are administered on a routine basis to supress lung infection [39]. The occurrence of both OCTT and SBWC accounting for significant explanation in both the whole microbiota and satellite, but not core taxa analysis, underpins the strong impact of gut physiology and transit on the microbiota in CF. Faecal calprotectin also explained the variance across the satellite taxa, and has been associated with increased abundances of *Escherichia, Streptococcus, Staphylococcus* and *Veillonella*, of which contained satellite species significantly increased in our CF group [26,40]. *Acidaminococcus sp*. have also associated with increased faecal calprotectin levels [28], with *Acidaminococcus intestinii* another constituent of the CF satellite microbiota that was not present in healthy controls (data not shown). The core taxa was only largely explained by the presence of CF disease itself, perhaps relating to the direct disruption of CFTR function which alone can influence changes in the microbiome [40].

Perhaps unsurprisingly, the species ordination biplots of the taxa from SIMPER analysis demonstrated clustering of the key SCFA producers mentioned previously away from the significant disease-associated clinical factors, with antibiotic usage and transit metrics previously shown to reduce the abundance of such taxa [14,41]. Similarly affected were taxa from genera that are associated with better outcomes in other similarly pro-inflammatory intestinal environments, such as Crohn’s disease or ulcerative colitis, including *Oscillibacter and Fusicanterbacter* [42,43].

*C. perfringens* has been associated with disease exacerbation in ulcerative colitis [36], SIBO in the CF mouse small intestine [44] and increased deconjugation of bile salts leading to further fat malabsorption by the host [45]. Here it was completely absent from our healthy control group, whilst in the CF group was found to associate with a variety of CF-induced clinical factors as well as OCTT. Also strongly associating with OCTT and impacted substantially more, was *E. coli*. Increased bacterial load relates to slower transit within the CF mouse small intestine [45]. Concurrently with the observed increase in SWBC reported prior [18], this in theory allows for the expansion of such facultative anaerobes in the small intestine that could potentially affect downstream community dynamics and functional profiles in the colon, given that PMA treatment was utilised to select for viable living taxa from faecal sampling.

Although dietary profiles were similar between groups (Tables S3-6) and did not contribute to significant variation in the microbiota, increased fat intake to meet energy requirements is a staple of the CF diet [46]. The infant gut metagenome demonstrates enrichment of fatty acid degradation genes [32] whilst CF-derived *E. coli* strains exhibit improved utilisation of exogenous glycerol as a growth source [47]. Finally, the genus *Bacteroides*, which has been reported to both increase and decrease within CF disease across different age groups [8,9,14], displayed high variability within the species ordination biplot (Fig. 3), perhaps resultant of the varying antimicrobial susceptibility within the genus [48].

We acknowledge the small sample size of this pilot study limits the power of specific analyses, with the absence of within-group direct ordination approaches which would have allowed for investigation of CF group antibiotic usage and extra clinical factors such as lung function. However, the principle strength of this study is the valuable insight into the relationships between microbiota composition and intestinal physiology and function in CF. Future studies should encompass larger cohorts in a longitudinal fashion with the combination of both lung and faecal microbiota data to elucidate such relationships better, including the impact of pulmonary antibiotic usage on the gut microbiota, and the aptly termed gut-lung axis. Evaluation of associations between the microbiota, physiology and the immune response would also improve our understanding of the mechanisms contributing to GI health in CF. Given their possible beneficial effect on intestinal inflammation [49], the impact of CFTR modulator therapy will provide further insights.

## 5. Conclusion

This cross-sectional pilot study has identified relationships between markers of clinical status, gastrointestinal function and bacterial dysbiosis in the CF population. By partitioning the community into core and satellite taxa, we were able to reveal the relative contributions of CF-associated lifestyle factors and elements of intestinal function to these subcommunity compositions, and how specific taxa were affected by these clinical factors. Further, as the first study to combine high-throughput gene amplicon sequencing with non-invasive MRI to assess underlying gut pathologies, we demonstrate the potential for future collaborations between gastroenterology and microbiology with larger cohort recruitment to investigate these relationships between gut function and the microbiome further.

## Data Availability

The data that support the findings of this study are openly available at figshare.com under https://doi.org/10.6084/m9.figshare.15073797.v1 and https://doi.org/10.6084/m9.figshare.15073797.v1 https://doi.org/10.6084/m9.figshare.15073899.v1 Raw sequence data reported in this study has been deposited in the European Nucleotide Archive under the study accession number PRJEB44071

https://doi.org/10.6084/m9.figshare.15073899.v1

## Author Contributions

CvdG, AS, GM, and RJM conceived the study. RJM, HG, LH, and MMR performed sample processing and analysis. RJM, DR, and CvdG performed the data and statistical analysis. CN, GM, and AS were responsible for sample collection, clinical care records and documentation. RJM, CN, GM and CvdG verified the underlying data. RJM, DR, and CvdG were responsible for the creation of the original draft of the manuscript. RJM, CN, GM, DR, AS, and CvdG contributed to the development of the final manuscript. CvdG is the guarantor of this work. All authors read and approved the final manuscript.

## Funding

A CF Trust Venture and Innovation Award (VIA 77) awarded to CvdG funded this work. Funding for the Gut Imaging for Function and Transit in CF (GIFT-CF) study was received from the Cystic Fibrosis Trust (VIA 061), Cystic Fibrosis Foundation (Clinical Pilot and Feasibility Award SMYTH18A0-I), and National Institute for Health Research Biomedical Research Centre, Nottingham.

## Declaration of Competing Interest

RJM, HG, LH, DM, and CvdG declare support from the CF Trust. CN and GM report grants and speaker honorarium from Vertex, outside the submitted work. ARS reports grants from Vertex, as well as speaker honoraria and expenses from Teva and Novartis and personal fees from Vertex, outside the submitted work. In addition, ARS has a patent issued “Alkyl quinolones as biomarkers of *Pseudomonas aeruginosa* infection and uses thereof”.

## Acknowledgements

We would like to thank Neele Dellschaft, Caroline Hoad, Luca Marciani, and Penny Gowland of the Sir Peter Mansfield Imaging Centre, University of Nottingham, who acquired the original MRI data [18]. We would also like to thank Nottingham Hospitals Charity, who funded faecal calprotectin assays and the transport of faecal samples for downstream analysis.

## Notes

### Author Declarations

West Midlands Coventry and Warwickshire Research Ethics Committee (18/WM/0242)

### Summary of Updates

Corrected Author Name: "Liam Hanson Hanson" changed to "Liam Hanson"

## References

[1] De Lisle RC, Borowitz D. The cystic fibrosis intestine. Cold Spring Harb Perspect Med 2013;3:a009753.

[2] Ooi CY, Durie PR. Cystic fibrosis from the gastroenterologist’s perspective. Nat Rev Gastroenterol Hepatol 2016;13:175–85.

[3] Tabori H, Arnold C, Jaudszus A, Mentzel HJ, Renz DM, Reinsch S, et al. Abdominal symptoms in cystic fibrosis and their relation to genotype, history, clinical and laboratory findings. PLoS One 2017;12:e0174463.

[4] Hayee B, Watson KL, Campbell S, Simpson A, Farrell E, Hutchings P, et al. A high prevalence of chronic gastrointestinal symptoms in adults with cystic fibrosis is detected using tools already validated in other GI disorders. United Eur Gastroenterol J 2019;7:881–8.

[5] Smith S, Rowbotham N, Davies G, Gathercole K, Collins SJ, Elliott Z, et al. How can we relieve gastrointestinal symptoms in people with cystic fibrosis? An international qualitative survey. BMJ Open Respir Res 2020;7:e000614.

[6] Antosca KM, Chernikova DA, Price CE, Ruoff KL, Li K, Guill MF, et al. Altered Stool Microbiota of Infants with Cystic Fibrosis Shows a Reduction in Genera Associated with Immune Programming from Birth. J Bacteriol 2019;201:e00274–19.

[7] Loman BR, Shrestha CL, Thompson R, Groner JA, Mejias A, Ruoff KL, et al. Age and environmental exposures influence the fecal bacteriome of young children with cystic fibrosis. Pediatr Pulmonol 2020;55:1661–70.

[8] Burke DG, Fouhy F, Harrison MJ, Rea MC, Cotter PD, Sullivan OO, et al. The altered gut microbiota in adults with cystic fibrosis. BMC Microbiol 2017;17:58.

[9] Vernocchi P, Chierico F Del, Russo A, Majo F, Rossitto M, Valerio M, et al. Gut microbiota signatures in cystic fibrosis: Loss of host CFTR function drives the microbiota enterophenotype. PLoS One 2018;13:e0208171.

[10] Kristensen M, Prevaes SMPJ, Kalkman G, Tramper-Stranders GA, Hasrat R, de Winter-de Groot KM, et al. Development of the gut microbiota in early life: The impact of cystic fibrosis and antibiotic treatment. J Cyst Fibros 2020.

[11] Donohoe DR, Garge N, Zhang X, Sun W, O’Connell TM, Bunger MK, et al. The Microbiome and Butyrate Regulate Energy Metabolism and Autophagy in the Mammalian Colon. Cell Metab 2011;13:517–26.

[12] Chang P V, Hao L, Offermanns S, Medzhitov R. The microbial metabolite butyrate regulates intestinal macrophage function via histone deacetylase inhibition. Proc Natl Acad Sci U S A 2014;111:2247–2252.

[13] Arpaia N, Campbell C, Fan X, Dikiy S, Veeken J van der, DeRoos P, et al. Metabolites produced by commensal bacteria promote peripheral regulatory T cell generation. Nature 2013;504:451–455.

[14] De Freitas MB, Moreira EAM, Tomio C, Moreno YMF, Daltoe FP, Barbosa E, et al. Altered intestinal microbiota composition, antibiotic therapy and intestinal inflammation in children and adolescents with cystic fibrosis. PLoS One 2018;13:e0198457.

[15] Flass T, Tong S, Frank DN, Wagner BD, Robertson CE, Kotter CV, et al. Intestinal lesions are associated with altered intestinal microbiome and are more frequent in children and young adults with cystic fibrosis and cirrhosis. PLoS One 2015;10:e0116967.

[16] Dayama G, Priya S, Niccum DE, Khoruts A, Blekhman R. Interactions between the gut microbiome and host gene regulation in cystic fibrosis. Genome Med 2020;12:12.

[17] Malagelada C, Bendezú RA, Seguí S, Vitrià J, Merino X, Nieto A, et al. Motor dysfunction of the gut in cystic fibrosis. Neurogastroenterol Motil 2020;00:e13883.

[18] Ng C, Dellschaft NS, Hoad CL, Marciani L, Ban L, Prayle AP, et al. Postprandial changes in gastrointestinal function and transit in cystic fibrosis assessed by Magnetic Resonance Imaging. J Cyst Fibros 2020.

[19] Gorzelak MA, Gill SK, Tasnim N, Ahmadi-Vand Z, Jay M, Gibson DL. Methods for improving human gut microbiome data by reducing variability through sample processing and storage of stool. PLoS One 2015;10:e0134802.

[20] Rogers GB, Cuthbertson L, Hoffman LR, Wing PAC, Pope C, Hooftman DAP, et al. Reducing bias in bacterial community analysis of lower respiratory infections. ISME J 2013;7:697–706.

[21] Callahan BJ, McMurdie PJ, Rosen MJ, Han AW, Johnson AJA, Holmes SP. DADA2: High resolution sample inference from Illumina amplicon data. Nat Methods 2016;13:4–5.

[22] Øyvind Hammer, David A.T. Harper and PDR. PAST: Paleontological Statistics Software Package for Education and Data Analysis. Palaeontol Electron 2001;4:9 p.

[23] ter Braak C, Smilauer P. CANOCO reference manual and user’s guide: software for ordination. Ithaca: Microcomputer Power; 2012.

[24] Cuthbertson L, Walker AW, Oliver AE, Rogers GB, Rivett DW, Hampton TH, et al. Lung function and microbiota diversity in cystic fibrosis. IthacaMicrobiome 2020;8:45.

[25] Zaneveld JR, McMinds R, Thurber RV. Stress and stability: Applying the Anna Karenina principle to animal microbiomes. Nat Microbiol 2017;2:17121.

[26] Enaud R, Hooks KB, Barre A, Barnetche T, Hubert C, Massot M, et al. Intestinal Inflammation in Children with Cystic Fibrosis Is Associated with Crohn’s-Like Microbiota Disturbances. J Clin Med 2019;8:645.

[27] Madan JC, Koestle DC, Stanton BA, Davidson L, Moulton LA, Housman ML, et al. Serial analysis of the gut and respiratory microbiome. MBio 2012;3:e00251–12.

[28] Coffey MJ, Nielsen S, Wemheuer B, Kaakoush NO, Garg M, Needham B, et al. Gut Microbiota in Children With Cystic Fibrosis: A Taxonomic and Functional Dysbiosis. Sci Rep 2019;9:18593.

[29] Al-Momani H, Perry A, Stewart CJ, Jones R, Krishnan A, Robertson AG, et al. Microbiological profiles of sputum and gastric juice aspirates in Cystic Fibrosis patients. Sci Rep 2016;6:26985.

[30] Anderson JL, Miles C, Tierney AC. Effect of probiotics on respiratory, gastrointestinal and nutritional outcomes in patients with cystic fibrosis: A systematic review. J Cyst Fibros 2017;16:186–97.

[31] Kastl AJ, Terry NA, Wu GD, Albenberg LG. The Structure and Function of the Human Small Intestinal Microbiota: Current Understanding and Future Directions. Cell Mol Gastroenterol Hepatol 2020;9:33–45.

[32] Manor O, Levy R, Pope CE, Hayden HS, Brittnacher MJ, Carr R, et al. Metagenomic evidence for taxonomic dysbiosis and functional imbalance in the gastrointestinal tracts of children with cystic fibrosis. Sci Rep 2016;6:22493.

[33] Hoffman LR, Pope CE, Hayden HS, Heltshe S, Levy R, McNamara S, et al. Escherichia coli dysbiosis correlates with gastrointestinal dysfunction in children with cystic fibrosis. Clin Infect Dis 2014;58:396–9.

[34] Hayden HS, Eng A, Pope CE, Brittnacher MJ, Vo AT, Weiss EJ, et al. Fecal dysbiosis in infants with cystic fibrosis is associated with early linear growth failure. Nat Med 2020;26:215–21.

[35] Duytschaever G, Huys G, Bekaert M, Boulanger L, Boeck K De, Vandamme P. Dysbiosis of bifidobacteria and Clostridium cluster XIVa in the cystic fibrosis fecal microbiota. J Cyst Fibros 2013;12:206–15.

[36] Li KY, Wang JL, Wei JP, Gao SY, Zhang YY, Wang LT, et al. Fecal microbiota in pouchitis and ulcerative colitis. World J Gastroenterol 2016;22:8929–39.

[37] Rainey FA, Hollen BJ, Small A. Genus I. Clostridium Prazmowski 1880, 23AL. 2nd ed. New York: Springer; 2009.

[38] Wang Y, Leong LEX, Keating RL, Kanno T, Abell GCJ, Mobegi FM, et al. Opportunistic bacteria confer the ability to ferment prebiotic starch in the adult cystic fibrosis gut. Gut Microbes 2019;10:367–81.

[39] Elborn JS. Cystic fibrosis. Lancet 2016;388:2519–31.

[40] Meeker SM, Mears KS, Sangwan N, Brittnacher MJ, Weiss EJ, Treuting PM, et al. CFTR dysregulation drives active selection of the gut microbiome. PLoS Pathog 2020;16:e1008251.

[41] Roager HM, Hansen LBS, Bahl MI, Frandsen HL, Carvalho V, Gøbel RJ, et al. Colonic transit time is related to bacterial metabolism and mucosal turnover in the gut. Nat Microbiol 2016;1:16093.

[42] Takeshita K, Mizuno S, Mikami Y, Sujino T, Saigusa K, Matsuoka K, et al. A single species of Clostridium Subcluster XIVa decreased in ulcerative colitis patients. Inflamm Bowel Dis 2016;22:2802–10.

[43] Vermeire S, Joossens M, Verbeke K, Wang J, Machiels K, Sabino J, et al. Donor Species Richness Determines Faecal Microbiota Transplantation Success in Inflammatory Bowel Disease. J Crohn’s Colitis 2016;10:387–94.

[44] Norkina O, Burnett TG, De Lisle RC. Bacterial overgrowth in the cystic fibrosis transmembrane conductance regulator null mouse small intestine. Infect Immun 2004;72:6040–9.

[45] De Lisle RC. Altered transit and bacterial overgrowth in the cystic fibrosis mouse small intestine. Am J Physiol - Gastrointest Liver Physiol 2007;293:104–11.

[46] Collins S. Nutritional management of cystic fibrosis – an update for the 21st century. Paediatr Respir Rev 2018;26:4–6.

[47] Matamouros S, Hayden HS, Hager KR, Brittnacher MJ, Lachance K, Weiss EJ, et al. Adaptation of commensal proliferating Escherichia coli to the intestinal tract of young children with cystic fibrosis. Proc Natl Acad Sci U S A 2018;115:1605–10.

[48] Nagy E, Urbán E, Nord CE. Antimicrobial susceptibility of Bacteroides fragilis group isolates in Europe: 20 years of experience. Clin Microbiol Infect 2011;17:371–9.

[49] Tetard C, Mittaine M, Bui S, Beaufils F, Maumus P, Fayon M, et al. Reduced Intestinal Inflammation With Lumacaftor/Ivacaftor in Adolescents With Cystic Fibrosis. J Pediatr Gastroenterol Nutr 2020;71:778–81.

