## Supplementary Materials for "Intestinal function and transit associate with gut microbiota dysbiosis in cystic fibrosis"

***Supplementary methods***

*Study participants and design*

Although 12 patients and controls were initially recruited, stool samples ultimately collected for 10 CF patients (6 males;4 females, Mean age of CF patients, 19.3 ± 7.93 years, Median age, 19 years) and of 10 non-CF healthy controls (Mean age of controls, 21.4 ± 7.40 years, Median age, 20 years). Patients were under a period of clinical stability and abstained from taking laxatives and anti-diarrhoeals during this visitation but were able to take routine pancreatic supplementation and perform standard physiotherapy procedures, with routine prophylactic antibiotic therapy recorded where applicable for patients.

*PMA treatment prior to DNA extraction*

1 mg PMA (Biotium, CA, USA) was hydrated in 98 µl 20% dimethyl sulfoxide (DMSO) to give a working stock concentration of 20 mM. 300 mg of stool thawed out from -80 °C was homogenised in 3 ml PBS, and centrifuged at 3200 g, for 5 minutes. The pellet was then resuspended in 1 ml prior to splitting into 500 µl fractions for subsequent PMA treatment. PMA 1.25 µl of PMA (20 mM) was added to give a final concentration of 50 µM. Following the addition of PMA to samples in opaque Eppendorf tubes, PMA was mixed by vortexing for 10 seconds, followed by incubation for 15 minutes at room temperature (~ 20 °C). This step was repeated before the transfer of samples to clear 1.5 mL Eppendorf tubes and placement within a LED lightbox. Treatment occurred for 15 minutes to allow PMA intercalation into DNA from compromised bacterial cells. Samples were then centrifuged at 10,000 x g for 5 minutes. The supernatant was discarded, and the cellular pellet was resuspended in 200 µl PBS.

*Targeted amplicon sequencing – Bacterial 16S rRNA*

Step 1 amplicon generation with primers based on the universal primer sequences 515F & 926R as described by Walters et al. (1), was performed under the following conditions; Initial denaturation of 180 seconds at 98 ºC, followed by: 25 cycles of 30 seconds at 95 ºC, 30 seconds at 55 ºC and 30 seconds at 72 ºC. A final extension of 5 minutes at 72 ºC was also included to complete the reaction. Step 2, the addition of dual barcodes and Illumina adaptor sequences was performed under the following conditions: Initial denaturation of 30 seconds at 98 ºC, followed by: 10 cycles of 10 seconds at 98 ºC, 20 seconds at 62 ºC and 30 seconds at 72 ºC. A final extension of 2 minutes at 72 ºC was also included to complete this reaction. This resulted in the generation of an ~ 550 bp amplicon spanning the V4-V5 hypervariable regions of the 16S rRNA gene.

*Sequencing Controls and Library Pooling*

PCR & DNA extraction negative controls were implemented, alongside the use of mock community positive controls, which included a Gut Microbiome Standard (ZYMO RESEARCH^TM^). Following Barcode attachment in the second PCR step, samples were normalised using the SequalPrep™ Normalization Plate Kit (Thermo Fisher Scientific), pooled and diluted to the final library concentrations requited for use on the Illumina MiSeq system.

*Sequence processing and analysis*

DADA2 was used to demultiplex and remove primer sequences, validate the quality profiles of forward and reverse reads and subsequently trim, infer sequence variants, merge denoised paired-reads, remove chimeras, and finally assign taxonomy via Naive Bayesian Classifier implementation. This included the use of the Genome Taxonomy Database (GTDB) reference sequences (2). Unidentifiable ASVs were run through a BLAST (<https://blast.ncbi.nlm.nih.gov/Blast.cgi>) and matched appropriately based on query coverage where possible. Taxa with 2 ≥ reads for a single sample were removed and excluded from subsequent statistical analysis. ASVs from the same bacterial taxon were collapsed to form a single OTU for a given taxon.

*References*

1.	Walters W, Hyde ER, Berg-lyons D, Ackermann G. Improved Bacterial 16S rRNA Gene (V4 and V4-5) and Fungal Internal Transcribed Spacer Marker Gene Primers for Microbial Community Surveys*. mSystems* 2016;1:e00009-15.

2.	Parks DH, Chuvochina M, Waite DW, Rinke C, Skarshewski A, Chaumeil PA, et al. A standardized bacterial taxonomy based on genome phylogeny substantially revises the tree of life. *Nat Biotechnol* 2018;36:996.

**Supplemntary Tables**

**Table S1** Dietary information obtained from study participants

| Study I.D. | Group | Mean % Kcal Protein | Mean % Kcal CHO | Mean % Kcal Fat | Mean % Fibre (g) |
| --- | --- | --- | --- | --- | --- |
| 152 | HC | 18.33 | 45.61 | 36.04 | 6.80 |
| 159 | HC | 18.16 | 46.96 | 32.91 | 4.46 |
| 205 | HC | 21.55 | 40.66 | 37.74 | 2.70 |
| 431 | HC | 8.46 | 63.62 | 27.98 | 3.38 |
| 501 | HC | 19.41 | 65.76 | 14.90 | 3.07 |
| 548 | HC | 16.55 | 43.20 | 34.01 | 6.28 |
| 673 | HC | 12.29 | 54.99 | 32.49 | 10.44 |
| 749 | HC | 14.70 | 42.82 | 36.14 | 6.79 |
| 964 | HC | 15.97 | 46.63 | 37.34 | 3.94 |
| 986 | HC | 15.88 | 43.83 | 40.25 | 4.88 |
| 128 | CF | 10.24 | 55.38 | 34.36 | 3.33 |
| 167 | CF | 17.38 | 44.21 | 38.59 | 5.07 |
| 259 | CF | 15.54 | 61.47 | 27.90 | 4.62 |
| 279 | CF | 11.97 | 47.36 | 40.41 | 2.11 |
| 297 | CF | 16.53 | 54.80 | 28.71 | 4.39 |
| 365 | CF | 17.19 | 49.28 | 33.51 | 4.21 |
| 596 | CF | 18.24 | 46.75 | 31.41 | 4.98 |
| 617 | CF | 12.15 | 51.63 | 32.89 | 4.07 |
| 643 | CF | 16.82 | 50.51 | 32.75 | 3.86 |
| 681 | CF | 15.46 | 50.70 | 33.74 | 3.47 |

Macronutrient intake 3 days prior to faecal sampling was recorded from participants through a food diary. The mean daily proportion of calories obtained from each macronutrient (Protein, CHO - Carbohydrates, Fat) was calculated. Mean relative weight of fibre intake was also calculated.

**Table S2** Magnetic resonance imaging (MRI) metrics utilised for the direct ordination approach.

| Study I.D. | Group (HC/CF) | OCTT (mins) | Corrected SBWC (mL/m^2^) | Corrected Fasting Colon Volume (mL/m^2^) |
| --- | --- | --- | --- | --- |
| 152 | HC | 180 | 61.51 | 755.63 |
| 159 | HC | 150 | 18.40 | 772.06 |
| 205 | HC | 360 | 40.35 | 708.42 |
| 431 | HC | 150 | 72.69 | 948.15 |
| 501 | HC | 360 | 47.56 | 393.86 |
| 548 | HC | 300 | 57.04 | 665.40 |
| 673 | HC | 360 | 24.16 | 985.48 |
| 749 | HC | 240 | 10.79 | 233.35 |
| 964 | HC | 180 | 26.19 | 649.25 |
| 986 | HC | 180 | 89.90 | 817.08 |
| 128 | CF | 150 | 55.44 | 435.61 |
| 167 | CF | 390 | 133.05 | 1389.02 |
| 259 | CF | 360 | 19.37 | 1048.02 |
| 279 | CF | 360 | 105.60 | 564.99 |
| 297 | CF | 180 | 273.09 | 845.09 |
| 365 | CF | 390 | 205.19 | 581.98 |
| 596 | CF | 120 | 83.36 | 646.69 |
| 617 | CF | 300 | 229.21 | 869.81 |
| 643 | CF | 390 | 82.92 | 719.90 |
| 681 | CF | 300 | 376.24 | 1667.85 |

HC - Healthy control, CF - Cystic fibrosis, OCTT – Oro-caecal transit time, SBWC – Small bowel water content corrected for body surface area, Colon Fasting Volume corrected for body surface area.

**Table S3** Summary statistics: Mean % Kcal Protein.

| Variable | Observations | Obs. with missing data | Obs. without missing data | Minimum | Maximum | Mean | Std. deviation |
| --- | --- | --- | --- | --- | --- | --- | --- |
| CF | 10 | 0 | 10 | 10.244 | 18.236 | 15.153 | 2.725 |
| HC | 10 | 0 | 10 | 8.464 | 21.552 | 16.131 | 3.726 |

Kruskal-Wallis test / Two-tailed test:

| K (Observed value) | 0.691 |
| --- | --- |
| K (Critical value) | 3.841 |
| DF | 1 |
| *P*-value (one-tailed) | 0.406 |
| alpha | 0.050 |

An approximation has been used to compute the *P*-value.

**Table S4** Summary statistics: Mean % Kcal CHO.

| Variable | Observations | Obs. with missing data | Obs. without missing data | Minimum | Maximum | Mean | Std. deviation |
| --- | --- | --- | --- | --- | --- | --- | --- |
| CF | 10 | 0 | 10 | 44.211 | 61.475 | 51.211 | 4.989 |
| HC | 10 | 0 | 10 | 40.664 | 65.757 | 49.409 | 8.932 |

Kruskal-Wallis test / Two-tailed test:

| K (Observed value) | 1.851 |
| --- | --- |
| K (Critical value) | 3.841 |
| DF | 1 |
| *P*-value (one-tailed) | 0.174 |
| alpha | 0.050 |

An approximation has been used to compute the *P*-value.

**Table S5** Summary statistics: Mean % Kcal Fat.

| Variable | Observations | Obs. with missing data | Obs. without missing data | Minimum | Maximum | Mean | Std. deviation |
| --- | --- | --- | --- | --- | --- | --- | --- |
| CF | 10 | 0 | 10 | 27.896 | 40.415 | 33.427 | 3.860 |
| HC | 10 | 0 | 10 | 14.902 | 40.254 | 32.981 | 7.203 |

Kruskal-Wallis test / Two-tailed test:

| K (Observed value) | 0.280 |
| --- | --- |
| K (Critical value) | 3.841 |
| DF | 1 |
| *P*-value (one-tailed) | 0.597 |
| alpha | 0.050 |

An approximation has been used to compute the *P*-value.

**Table S6** Summary statistics: Mean % Fibre (g).

| Variable | Observations | Obs. with missing data | Obs. without missing data | Minimum | Maximum | Mean | Std. deviation |
| --- | --- | --- | --- | --- | --- | --- | --- |
| CF | 10 | 0 | 10 | 2.106 | 5.070 | 4.010 | 0.883 |
| HC | 10 | 0 | 10 | 2.703 | 10.438 | 5.274 | 2.356 |

Kruskal-Wallis test / Two-tailed test:

| K (Observed value) | 0.966 |
| --- | --- |
| K (Critical value) | 3.841 |
| DF | 1 |
| p-value (one-tailed) | 0.326 |
| alpha | 0.050 |

An approximation has been used to compute the *P*-value.

**Table S7** Core taxa within each group throughout the study.

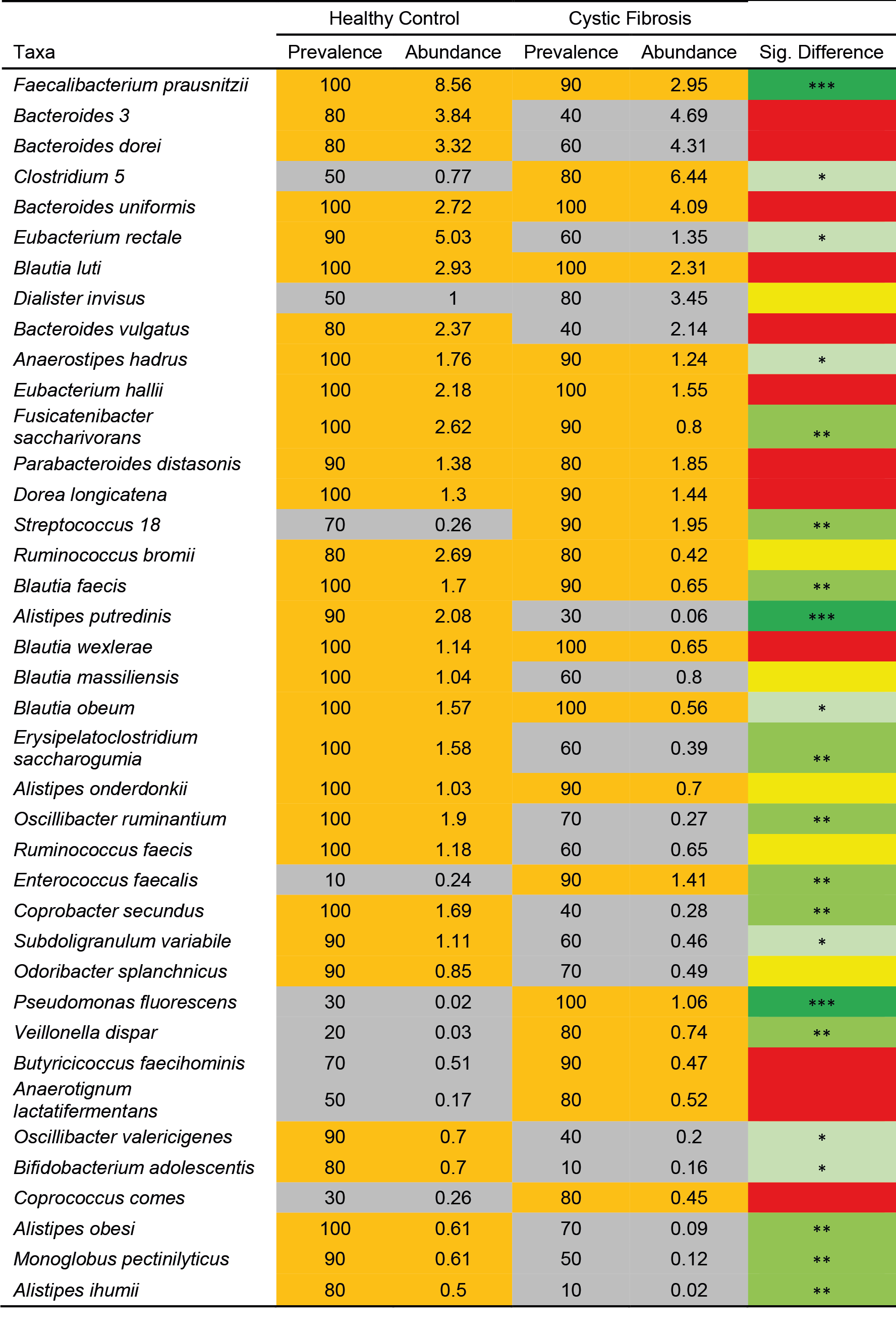

Given is prevalence, the percent number of samples a given core taxon was detected in, and average relative abundance across those samples. Taxon names are derived from condensed ASVs of the same species. ASV numbers have been used to differentiate between taxa within the same genus that could not be identified at the species level. Core taxa are highlighted orange, whereas satellite taxa are grey. Given the length of the ribosomal sequences analysed, species identities should be considered putative. *** - *p* < 0.001, ** - *p* < 0.01, * - *p* < 0.05.

**Table S8** Bacterial Kruskal-Wallis tests of alpha diversity.

| **Microbiota** |  | **Core taxa** |  | **Satellite taxa** |  |
| --- | --- | --- | --- | --- | --- |
| H: | 7.406 | H: | 14.29 | H: | 7 |
| Hc (tie corrected): | 7.406 | Hc (tie corrected): | 14.29 | Hc (tie corrected): | 7 |
| *p* (same): | **0.006502** | *p* (same): | **0.000157** | *p* (same): | **0.008151** |

**Table S9** Bacterial ANOSIM summary statistics utilising Bray-Curtis index.

| **Microbiota** |  | **Core taxa** |  | **Satellite taxa** |  |
| --- | --- | --- | --- | --- | --- |
| R value: | 0.4729 | R value: | 0.9022 | R value: | 0.5609 |
| *p* (same): | **0.0001** | *p* (same): | **0.0001** | *p* (same): | **0.0001** |
| Bonferroni-corrected p value: | 0.0001 | Bonferroni-corrected p value: | 0.0001 | Bonferroni-corrected p value: | 0.0001 |
| permutation *N*: | 9999 | permutation *N*: | 9999 | permutation *N*: | 9999 |
